# Excess deaths due to incident Alzheimer’s dementia in women and men in the United States

**DOI:** 10.1101/2025.08.27.25334590

**Authors:** Bryan D. James, Tianhao Wang, S.E. Leurgans, Lisa L. Barnes, David X. Marquez, David A. Bennett

**Author notes:** Correspondence: Bryan D. James, Rush Alzheimer’s Disease Center, Rush University Medical Center, Chicago, IL 60612, USA. Funding information: National Institutes of Health, Grant/Award Numbers: R01AG017917, P30AG072975, R01AG022018.

## Abstract

**Objectives:** To assess whether the burden of mortality attributable to Alzheimer’s dementia in the US for women and men.

**Methods:** Data came from 3,491 women and 1,160 men ages 65 and older (mean 76.5 for both sexes) with no dementia at baseline from five longitudinal cohort studies of aging with identical annual diagnostic assessments of dementia. Mortality hazard ratios (HR) after incident Alzheimer’s dementia were estimated per 10-year age strata from proportional hazard models. Population attributable risk percent (PAR%) was derived to estimate excess mortality after a diagnosis of incident Alzheimer’s dementia. Results were then stratified by self-reported sex, and separately with an interaction term for sex by incident Alzheimer’s dementia. The number of excess deaths attributable to Alzheimer’s dementia in the US for women and men by age group was then estimated.

**Results:** Over an average of 9 (SD=5.8) years, 954 (27.3%) women and 316 (27.2%) men without dementia at baseline developed Alzheimer’s dementia; 1,792 (51.3%) women and 726 (62.6%) men died. In a model with terms for sex, race, education, incident Alzheimer’s dementia, and interaction between male sex and Alzheimer’s dementia, we observed an interaction (HR = 1.24, 95% CI: 1.00, 1.53) in the age strata 85+, indicating a higher risk of mortality due to Alzheimer’s dementia for men; at lower ages the interaction was opposite (HR = 0.75, 95% CI: 0.52, 1.09 in age strata 75-84), indicating higher risk of mortality from Alzheimer’s dementia for females. After further adjusting for vascular risk factors and diseases, the interactions were similar. PAR% was similar for age 85+ for women and men (33.4% and 32.9% respectively) but higher for women than men in the age strata 75-84 (24.2% and 19.1%). In 2023, we estimate 465,400 deaths—271,700 in women and 193,700 in men—were attributable to Alzheimer’s dementia. Adjusted PAR%s that took account of differences in vascular risk factors and disease showed even larger gaps for women compared to men (41.3% vs 37.5% for age 85+ and 33.2% vs 16.3% for age 75-84), resulting in estimates of 349,409 deaths from AD for women and 198,724 for men.

**Conclusions:** The number of deaths attributable to Alzheimer’s dementia is estimated to be 270,000-350,000, making it one of the leading causes of death in women, on par with cancer. The number is about 200,000 in men which also makes it a leading cause of death, on a par with accidents but much lower than cancer.

## INTRODUCTION

Alzheimer disease (AD) is listed by the Centers for Disease Control and Prevention (CDC) as the sixth leading cause of death in the United States (114,034 deaths in 2023); broken down by sex, the CDC lists it as the 4^th^ leading cause of death for women (77, 974 or 46.1 per 100,000 women) and 8^th^ leading cause of death for men (36,060 or 21.8 per 100,000 men) [1]. However, CDC causes of death are from death certificates which largely reflect proximate cause of death rather than underlying cause of death.

Further, the CDC reports a single cause of death for each person, which was more appropriate when infectious diseases were the primary cause of death. However, since the middle to late 20^th^ century, death is much more frequently the result of multi-morbidity, especially for older adults [2, 3]. Another approach to examining the contribution of a chronic disease like Alzheimer’s dementia that does not rely on a single cause of death is to estimate the attributable risk of death due to Alzheimer’s dementia [4–6].

A previous study by our group using the attributable risk approach estimated about 500,000 deaths attributable to incident Alzheimer’s dementia [7]. However, we were unable to generate sex-specific differences. The population attributable risk approach used in this previous study does not rely upon what is written on death certificates and instead is calculated based on observed mortality rates as the proportion of deaths in persons who develop Alzheimer’s dementia in excess of deaths in persons who do not. Here we extend this approach to examine the mortality risk for Alzheimer’s dementia and excess fraction of deaths due to Alzheimer’s dementia separately for women and men, by age group. We used data from five community-based cohort studies of older adults with harmonized annual cognitive assessments and diagnostic criteria to compare the following estimates for women and men: the age group-specific mortality hazard ratio for Alzheimer’s dementia, the population attributable risk for Alzheimer’s dementia, and the number of death attributable to Alzheimer’s dementia.

## METHODS

### Participants

Data comes from participants enrolled in five ongoing prospective cohorts at RADC with harmonized cognitive batteries, allowing them to be pooled for thee analyses[8]. The Religious Orders Study (ROS) was initiated in 1994 and enrolls older Catholic nuns, priests, and brothers from across the United States.[9] The Rush Memory and Aging Project (MAP) started in 1997 and enrolls older adults from the Chicago metropolitan area.[9] The Minority Aging Research Study (MARS)[10] was started in 2004, and the Rush Clinical Core[11] was started in 2008; both studies recruit older African Americans from churches, senior buildings, and social clubs and organizations that serve diverse populations in the Chicago metropolitan area. The Rush Latino Core launched in 2015 and includes older adults in Chicago who self-identify as Latino.[8] All five cohort studies require annual cognitive testing. Follow-up rate was 85% to 90% in these cohorts. All studies were approved by an institutional review board of Rush University Medical Center. All participants signed informed consent and ROS and MAP required a signed Anatomic Gift Act for brain donation; brain donation is optional in the other three cohorts.

### Clinical evaluation and diagnosis of AD dementia

Annual clinical evaluations included medical history, neurologic examination, and cognitive testing.[8, 12] The medical history includes questions regarding vascular disease history (e.g., claudication, stroke, and heart conditions) and vascular risk factors (hypertension, diabetes mellitus, and smoking); each were summed for analysis, range: 0–3. Clinical diagnosis of Alzheimer’s and other dementias at each assessment was performed using a 3-stage process with computer scoring of cognitive tests followed by clinical judgment by a neuropsychologist, and diagnostic classification by an experienced clinician.[12–14] Diagnosis of dementia and Alzheimer’s dementia followed National Institute of Neurological and Communicative Disorders and Stroke–Alzheimer’s Disease and Related Disorders Association criteria.[14] The diagnosis of Alzheimer’s dementia requires evidence of a meaningful decline in cognitive function relative to a previous level of performance with impairment in memory and at least one other area of cognition.

### Ascertainment of mortality

In addition to the annual clinical evaluations, participants are contacted regularly to determine vital status. When a participant cannot be located, we call one on the contacts provided and death is occasionally reported by the contact. Finally, the Social Security Death Index is regularly searched for the small number of participants we are unable to contact anyone. Furthermore, as all cohorts either require (ROS and MAP) organ donation at death or make the option to donate available, death is closely monitored and family members alert study staff when participants are near death or pass as soon as possible. Thus, for nearly all deceased participants, the exact day of death is known.

### Statistical Analysis

#### Survival after Alzheimer’s dementia diagnosis for men and women

We first compared median survival time overall and at three age strata (65–74 years, 75–84 years, and 85 years and older) for men and women. Kaplan-Meier curves were obtained for the three age strata to estimate the median time to death after a diagnosis of Alzheimer’s dementia separately for men and women.

#### Risk of mortality from incident Alzheimer’s dementia for men and women

Proportional hazards models with age as the time scale were used to calculate mortality hazard ratios (HRs) for incident Alzheimer’s dementia for the three age strata. Participants without dementia entered the analysis at baseline age (left truncation). Alzheimer’s dementia status was treated as a time-varying absorbing state; person-years for subjects who developed Alzheimer’s dementia contributed to mortality hazard for no Alzheimer’s dementia up to diagnosis, and then to mortality hazard for Alzheimer’s dementia after diagnosis. Models included terms for sex, as well as race/ethnicity (terms for Black race and Latino ethnicity with non-Latino white as reference) and education. We then constructed a model with an interaction term for sex times Alzheimer’s dementia to characterize the potential sex difference in the association of incident Alzheimer’s dementia with mortality. A third model also included terms for vascular risk factors and disease burden. We repeated these models with a term for all cause dementia in place of Alzheimer’s dementia.

#### Population attributable risk percentage: Excess deaths due to incident Alzheimer’s dementia for men and women

Population attributable risk percentage (PAR%), also known as “excess fraction”, is calculated as the proportion of new cases of an outcome that occurs in the exposed group in excess of new cases in the unexposed group. [7, 15] In this analysis, PAR% is an estimate of total mortality risk that can be considered attributable to developing Alzheimer’s dementia; i.e., the proportion of deaths that occur after developing Alzheimer’s dementia that is in excess of deaths among people without Alzheimer’s dementia. The crude PAR% is estimated by:

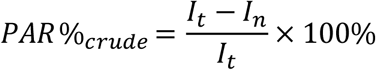

where *I*_*t*_ is the mortality rate of the entire cohort, and *I*_*n*_ is the mortality rate among the person-years without Alzheimer’s dementia.

To calculate adjusted PAR% using a mortality hazard ratio adjusted for covariates (r), the prevalence proportion of Alzheimer’s dementia (p) is necessary:

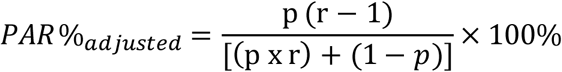

Because prevalence cannot be accurately calculated based on the design of the cohorts used for this study in which persons with known dementia were excluded, we used sex- and age-specific US prevalence estimates from the population-based Health and Retirement Study (HRS).[16] Prevalence estimates (as proportions) were as follows: men ages 65-74: 2.27%, ages 75-84: 7.79%, age 85+: 25.84; women ages 65-74: 3.75%, ages 75-84%: 12.05, age 85+: 41.26%. While these estimates were for the year 2012, they were the most recent sex- and age-specific prevalence estimates for the US available.

#### Number of deaths attributable to Alzheimer’s dementia in the United States for men and women

Finally, to compare the actual predicted number of men and women in the US for which Alzheimer’s dementia contributed to their deaths, we applied the age- and sex-specific PAR% estimates to the reported numbers of total US deaths in these age ranges by sex for 2025 as projected from the US Census.[1]

## RESULTS

Analyses included 3,491 women and 1,160 men age 65 and older (mean age 76.5 for both sexes) who were followed for an average of 9 (SD=5.8) years. The cohort was 64% non-Latino White, 27% Black, and 9% Latino (less than 1% “other” or race/ethnicity not recorded); mean education was 15.5 years (SD=3.8). During follow-up, 954 (27.3%) women and 316 (27.2%) men developed Alzheimer’s dementia, and 1,792 (51.3%) women and 726 (62.6%) men died.

### Median survival time after Alzheimer’s dementia diagnosis for men and women

The median survival time from Alzheimer’s dementia diagnosis to death from Kaplan-Meier curves for participants who developed Alzheimer’s dementia was 3.4 years overall: 4.3 years for persons aged 75–84 at Alzheimer’s dementia diagnosis (n = 293) and 2.7 for persons aged 85 and older at Alzheimer’s dementia diagnosis (n = 677); only 32 persons aged 65–74 died after developing Alzheimer’s dementia, so Kaplan-Meier curve could not be estimated. Survival after diagnosis was 3.6 years for women and 2.7 for men. As shown in Figure 1, among persons aged 75–84 at Alzheimer’s dementia diagnosis, median survival was 5.1 years for females (n = 206) and 3.8 for males (n = 87). Among persons aged 85 and older at Alzheimer’s dementia diagnosis, median survival was 3.0 years for females (n = 509) and 1.9 for males (n = 168).

**Figure 1.**
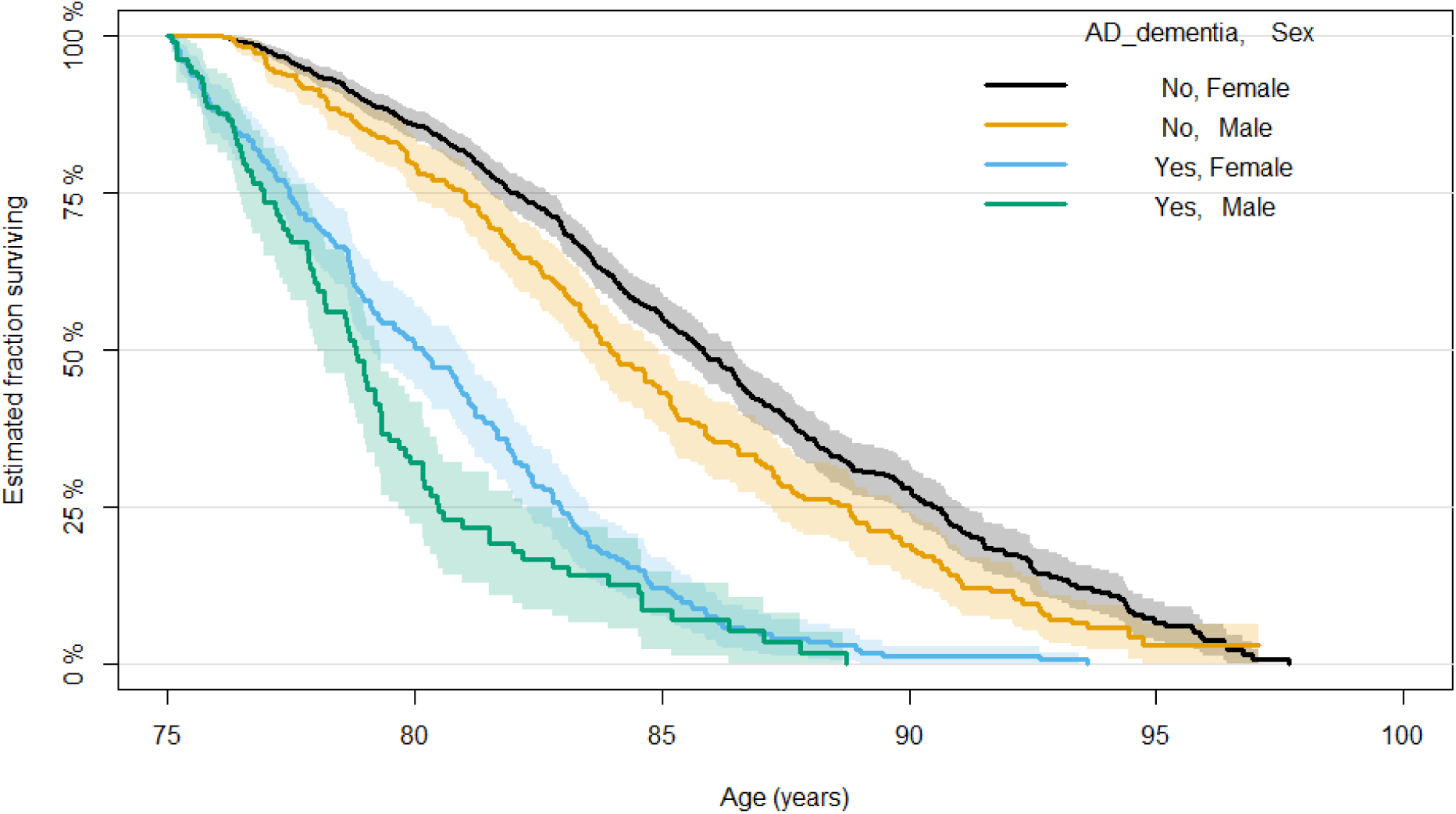
Survival after a diagnosis of Alzheimer’s dementia vs no diagnosis of Alzheimer’s dementia for men and women, ages 75-84 and 85+. **Figure 1a. Age 75-84.**

**Figure 1b.**
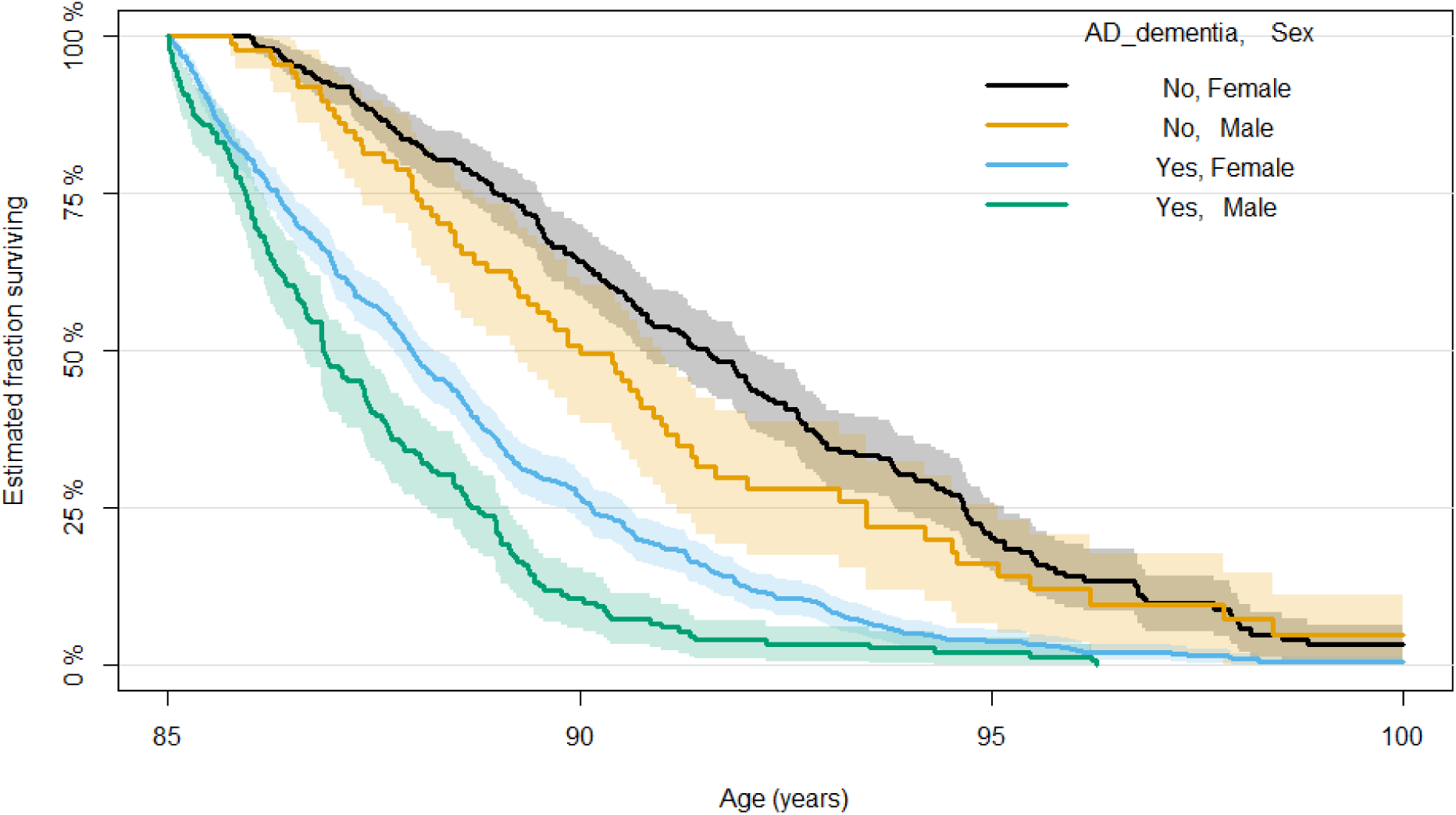
Age 85+.

### Risk of mortality due to developing Alzheimer’s dementia for men and women

In proportional hazards models model with terms for sex, race/ ethnicity, and education, incident Alzheimer’s dementia was strongly associated with mortality across age groups, though the association attenuated from a HR of 5.2 for age 65-74 to 2.9 for age 85 and older (Table 1a). In models that included a term for the interaction between male sex and Alzheimer’s dementia, we observed an interaction (HR = 1.24, 95% CI: 1.00, 1.53) in the age strata 85+, indicating a higher risk of mortality due to Alzheimer’s dementia for males compared to females; however, at lower ages the interaction was opposite, indicating higher risk of mortality from Alzheimer’s dementia for females, though not statistically significant (Table 1b). Figure 2 displays the estimated mortality HR for Alzheimer’s dementia for women and men for ages 75-84 and 85+ (ages 65-74 not displayed due to small number of deaths after developing Alzheimer’s dementia). The risk of death was much higher for women in the younger age strata compared to age 85+, but similar for men across the two age strata. After further adjusting for vascular risk factors and diseases, the interactions were similar, although the interaction indicating higher risk of mortality from Alzheimer’s dementia for females (i.e., lower for males) was significant for ages 75-84 (HR = 0.68, 95% CI: 0.47, 0.99). Estimates were very similar with the same inferences when rerunning models after replacing Alzheimer’s dementia with dementia of any type.

**Figure 2.**
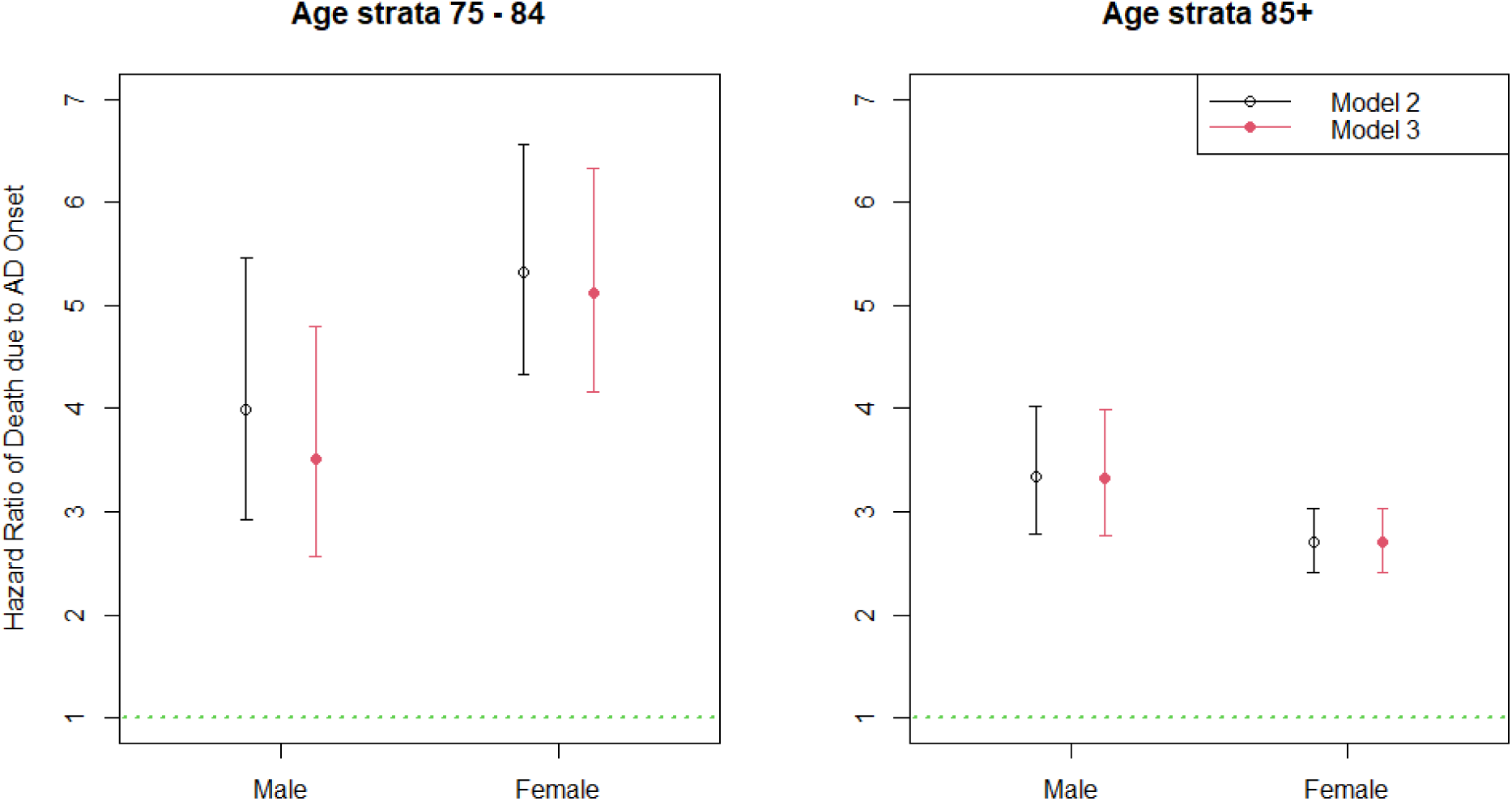
Hazard ratio of death due to Alzheimer’s dementia onset for men and women, age 75-84 and age 85+. Model 2: includes terms for incident Alzheimer’s dementia, sex, race/ethnicity (terms for Black race and Latino ethnicity with non-Latino white as reference), education and the interaction of sex and Alzheimer’s dementia. Model 3: Terms from Model 2 as well as terms for vascular risk factors and disease burden

**Table 1a.** Cox model without interaction between Alzheimer’s dementia and sex.

|  | Age<br>65-74 |  |  |  | Age<br>75-84 |  |  |  | Age<br>85+ |  |  |  |
| --- | --- | --- | --- | --- | --- | --- | --- | --- | --- | --- | --- | --- |
|  | HR | lower<br>.95 | upper<br>.95 | p-<br>value | HR | lower<br>.95 | upper<br>.95 | p-<br>value | HR | lower<br>.95 | upper<br>.95 | p-<br>value |
| **Male sex** | 1.54 | 1.04 | 2.29 | 0.032 | 1.51 | 1.28 | 1.79 | 0.000 | 1.49 | 1.34 | 1.66 | 0.000 |
| **Incident**<br>**Alzheimer's**<br>**dementia** | 5.18 | 2.68 | 9.99 | 0.000 | 4.86 | 4.09 | 5.79 | 0.000 | 2.86 | 2.60 | 3.16 | 0.000 |
| **Black race** | 2.15 | 1.48 | 3.11 | 0.000 | 1.17 | 0.99 | 1.39 | 0.067 | 0.90 | 0.77 | 1.05 | 0.163 |
| **Latino** | 1.00 | 0.49 | 2.04 | 0.991 | 0.87 | 0.60 | 1.25 | 0.437 | 0.98 | 0.74 | 1.29 | 0.865 |
| **Education** | 1.04 | 1.01 | 1.07 | 0.016 | 1.01 | 0.99 | 1.02 | 0.444 | 0.99 | 0.98 | 1.00 | 0.051 |

**Table 1b.**
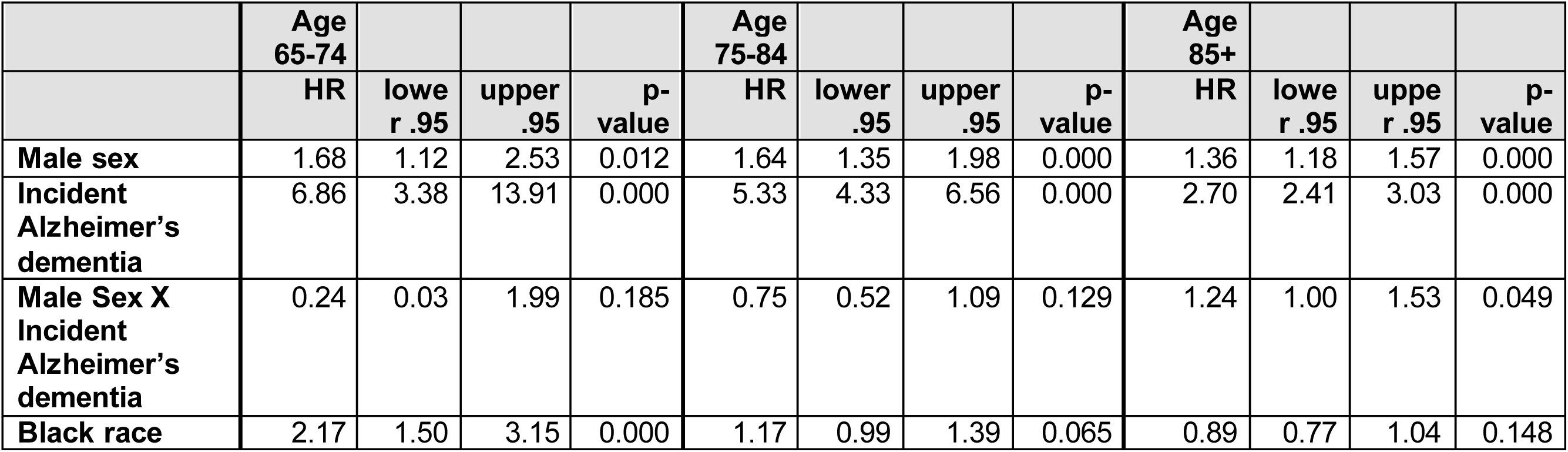
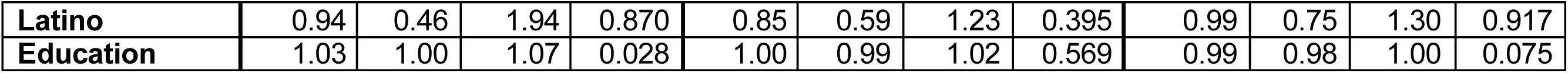
Cox model with interaction between AD and sex.

|  | Age<br>65-74 |  |  |  | Age<br>75-84 |  |  |  | Age<br>85+ |  |  |  |
| --- | --- | --- | --- | --- | --- | --- | --- | --- | --- | --- | --- | --- |
|  | HR | lower<br>.95 | upper<br>.95 | p-<br>value | HR | lower<br>.95 | upper<br>.95 | p-<br>value | HR | lower<br>.95 | upper<br>.95 | p-<br>value |
| **Male sex** | 1.68 | 1.12 | 2.53 | 0.012 | 1.64 | 1.35 | 1.98 | 0.000 | 1.36 | 1.18 | 1.57 | 0.000 |
| **Incident**<br>**Alzheimer's**<br>**dementia** | 6.86 | 3.38 | 13.91 | 0.000 | 5.33 | 4.33 | 6.56 | 0.000 | 2.70 | 2.41 | 3.03 | 0.000 |
| **Male Sex X**<br>**Incident**<br>**Alzheimer's**<br>**dementia** | 0.24 | 0.03 | 1.99 | 0.185 | 0.75 | 0.52 | 1.09 | 0.129 | 1.24 | 1.00 | 1.53 | 0.049 |
| **Black race** | 2.17 | 1.50 | 3.15 | 0.000 | 1.17 | 0.99 | 1.39 | 0.065 | 0.89 | 0.77 | 1.04 | 0.148 |
| **Latino** | 0.94 | 0.46 | 1.94 | 0.870 | 0.85 | 0.59 | 1.23 | 0.395 | 0.99 | 0.75 | 1.30 | 0.917 |
| **Education** | 1.03 | 1.00 | 1.07 | 0.028 | 1.00 | 0.99 | 1.02 | 0.569 | 0.99 | 0.98 | 1.00 | 0.075 |

**Table 1c.**
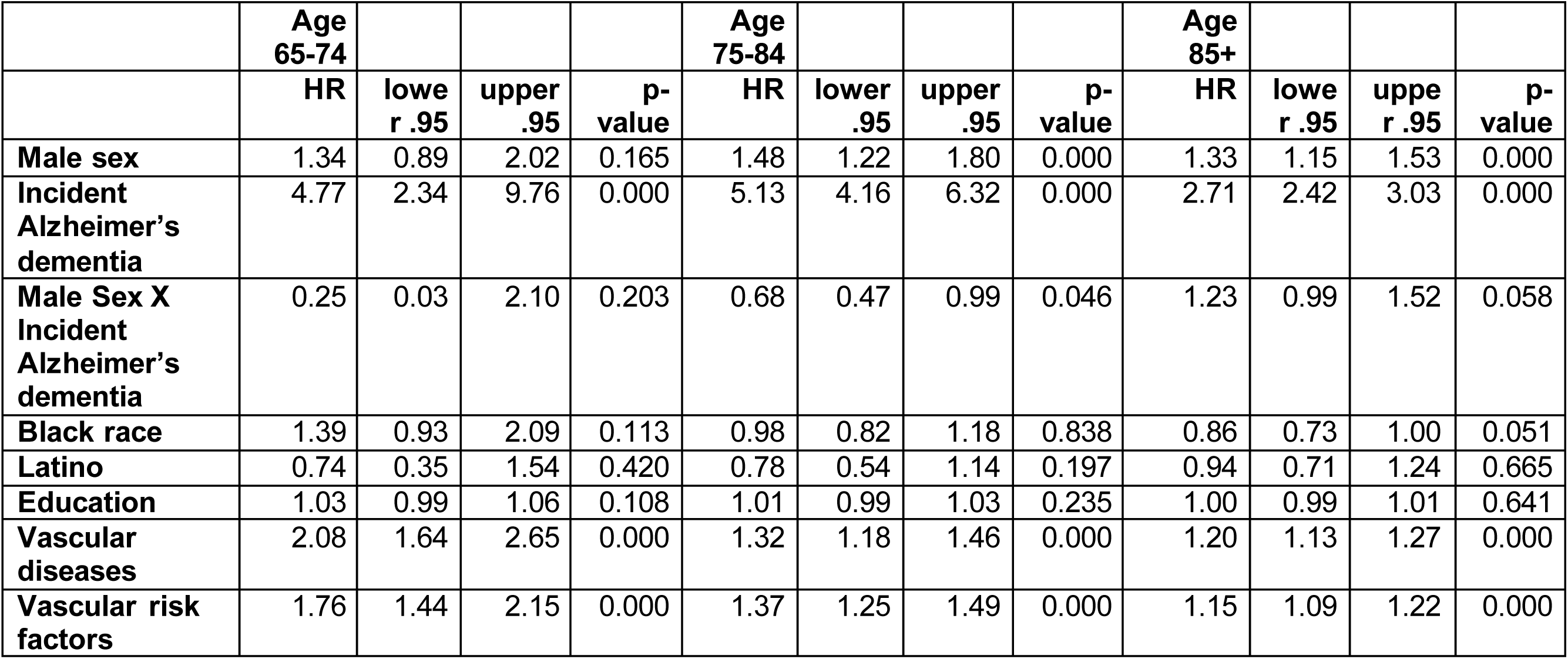
Cox model adjusting for vascular diseases and risks.

### Population attributable risk percentage (PAR%): Excess deaths due to Alzheimer’s dementia for women and men

We then calculated PAR% separately for women and men using the sex- and age-specific HR estimates from the previous models. In the age strata 75-84, PAR% was 24.2% for women and 19.1% for men. By contrast, in the age strata 85+ it was 33.4% for women and 32.9% for men (Table 2). Age- and sex-specific PAR% was slightly higher though very similar for dementia of any type (25.3% and 21.1%, and 36.5% and 34.6% respectively), so we will focus on Alzheimer’s dementia throughout.

**Table 2.** Sex- and Age-Specific Population Attributable Risk Percentages Female.

| Age groups | No Alzheimer's dementia |  |  | Alzheimer's dementia |  |  | PAR% |
| --- | --- | --- | --- | --- | --- | --- | --- |
|  | # deaths | Person-years | Mortality rate* | # deaths | Person-years | Mortality rate* |  |
| **65-74** | 71 | 7,274.3 | 9.8 | 7 | 101.8 | 68.8 | --** |
| **75-84** | 297 | 12,930.1 | 23.0 | 121 | 871.1 | 138.9 | 24.2% |
| **85+** | 614 | 7,984.7 | 76.9 | 547 | 2,077.5 | 263.3 | 33.4% |

**Male**
| Age groups | No Alzheimer's dementia |  |  | Alzheimer's dementia |  |  | PAR% |
| --- | --- | --- | --- | --- | --- | --- | --- |
|  | # deaths | Person-years | Mortality rate* | # deaths | Person-years | Mortality rate* |  |
| **65-74** | 34 | 2,383.5 | 14.3 | 1 | 29.0 | 34.5 | --** |
| **75-84** | 155 | 4,295.3 | 36.1 | 50 | 301.4 | 165.9 | 19.1% |
| **85+** | 251 | 2,516.5 | 99.7 | 195 | 484.5 | 402.5 | 32.9% |

\* Per 1000 person-years
\*\* Because of the small number of deaths in men and women age 65-74 who developed Alzheimer's dementia, we do not present PAR% estimates for this age range

We then calculated adjusted PAR%, based on HRs from proportional hazards models adjusted for sex, race/ ethnicity, education, and vascular risk factors and diseases as well as age-specific prevalence rates adapted from HRS[16]. For women, the adjusted PAR% was 33.2% and 41.3% for age strata 75-84 and 85+, respectively. By contrast, it was 16.3% and 37.5% and for men in those age strata.

### Number of deaths attributable to Alzheimer’s dementia in the United States for men and women

By applying sex- and age-specific estimates of crude PAR% to the numbers of deaths in women and men in the United States in 10-year age groups[US Census], we estimated 271,700 deaths in women (93,450 in ages 75-84; 178,450 in ages 85+) and 193,700 deaths in men (78,550 in ages 75-84; 115,150 in ages 85+) were attributable to incident Alzheimer’s dementia in 2023 (the most recent year census data is available). Data was insufficient to estimate this number for persons younger than age 75. When using the adjusted PAR% instead, the number of deaths from Alzheimer’s dementia was much higher for women (349,409), while the estimated number of deaths from Alzheimer’s dementia for men was similar to the estimate using crude PAR% (198,724).

## DISCUSSION

In this prospective study, older women and men from five harmonized cohort studies were followed for an average of 9 years to assess the risk of mortality after developing dementia at different ages, and to estimate the proportion and number of deaths in women and men that can be attributed to Alzheimer’s dementia in the U.S. We found that the overall proportion and number of deaths that can be attributed to Alzheimer’s dementia in women was about 270,000; the estimate increased to about 350,000 with the adjusted PAR% which takes prevalence into account. By contrast, about 200,000 were attributable to Alzheimer’s dementia in men. These numbers are several fold higher than reports from the CDC based on death certificates. Direct comparisons cannot be made to death certificate reporting which reports a single cause of death that is much more likely to be the proximate cause of death. However, our findings suggest that Alzheimer’s is a much more significant contributor to death for women than generally recognized, with a death toll on par with heart disease and cancer, the number one and two killers of American women.[1] The number of deaths attributable to Alzheimer’s dementia for men was also much higher than what is reported on death certificates, but similar to accidents and much less than for heart disease and cancer.

Incident Alzheimer’s dementia was strongly related to mortality risk for both women and men and the hazard ratio attenuated with higher age for both sexes; however, we observed evidence of qualitative interactions for this relationship by sex across age groups: while the risk of mortality due to Alzheimer’s dementia stayed relatively constant across age strata in males, it was much higher for females age 75-84 and lower for females age 85+. Across all age strata, women lived longer after developing Alzheimer’s dementia than men. It is difficult to directly compare these results to previous research that used different methodology or study populations, but results have been equivocal regarding sex differences in risk of mortality from Alzheimer’s dementia. In death certificate studies, mortality is usually reported as higher for women. For example, in U.S. vital statistics, women have higher age-adjusted mortality rates than men [1, 17]. A multi-country analysis of European vital statistics similarly found higher age-adjusted Alzheimer’s mortality in women from 2012-2010 [18]. A study in Australia reported the same, however they found that women’s older age at death explained much of the sex-related difference in rates of death from dementia, though some excess for deaths from Alzheimer’s dementia specifically did remain in women [19]

By contrast, studies that do not rely upon death certificate information report the opposite of death certificate studies: a higher rate of mortality from Alzheimer’s dementia in men. A recent nationwide study of US Medicare enrollees that compared survival in men and women with codes for dementia in their claims reported 24% higher adjusted mortality in males with dementia [20] A population-based analysis of HRS data similarly found the association between dementia and earlier death was stronger in men than women. [21] This could indicate biases by sex on the accuracy or sensitivity of Alzheimer’s death reporting in death certificates, but more research is needed to determine whether differences in measurement, methodology, geographic location or ways of accounting for differences at different ages account for these differences in findings. Our findings show that simply adjusting for age may mask sex-differences in risk of mortality from Alzheimer’s dementia, which may be higher in women at younger ages but lower at higher ages. We are aware of only other study that prospectively estimated the contribution of Alzheimer’s dementia to mortality for men and women in this fashion.[22] Interestingly, it reported that over 15 years of follow up, Alzheimer’s dementia increased mortality overall, but this association was significant only for women and not for men. Duration of survival from incident Alzheimer’s to death was longer in women than men in the age 75-84 and 85+ strata, similar to the findings here. Unfortunately, population attributable risk was only reported for the entire cohort and not for women and men separately.

When extrapolating our population attributable risk proportion estimate to the number of deaths in US women and men age 75 and older, the number of deaths attributable to Alzheimer’s dementia in women was about 40% higher than the number in men; at age 85 and older, the number of deaths attributable to Alzheimer’s was 55% higher for women. Thus, while the risk of death after developing Alzheimer’s dementia is higher for men than women for persons age 85 and older, Alzheimer’s dementia still proportionately contributes to more of the total deaths in women in this age range.

Interestingly, these ratios are very similar to the ratios observed when comparing Alzheimer’s disease death rates per 100,000 for women and men as reported by the CDC: 40% higher for age 75-84 (220.6 for women compared to 157.9 for men) and 51% higher for age 85+ (1,259.3 for women compared to 832.0 for men).[1].

When we calculated adjusted PAR% from hazard ratios that adjusted for differences in vascular diseases and risk factors and took into account the higher prevalence of Alzheimer’s dementia in US women, the gaps in attributable risk between women and men were even higher. Crude and adjusted PAR% were very similar for men, but adjusted PAR% was much higher for women, resulting in an estimate of about 350,000 deaths attributable to Alzheimer’s in women. More work is required to understand if this widening gap in attributable risk when adjusting for vascular risk factors and diseases is supportive of the argument that the observed higher burden of Alzheimer’s dementia in women is due to survival bias from cardiovascular disease; i.e., that men are more likely to die from cardiovascular disease in middle age and thus the ones that survive to be observed at older ages have healthier cardiovascular risk profiles and thus lower risk for dementia.[23, 24] Alternately, men diagnosed with dementia often have more comorbid conditions, which can raise post-diagnosis mortality.[25]

This study has limitations, including the fact that there were three times as many female participants as male, which could lead to more accurate estimates in women than men. Additionally, we did not observe enough deaths after developing Alzheimer’s dementia in the younger ages to provide an accurate estimate of PAR% in the 65-74 age group.

Another limitation was the inability to calculate prevalence estimates from the cohort studies used for the hazard ratio estimates, thus relying on published sex- and age-specific prevalence estimates from HRS, which utilizes alternate dementia ascertainment methods. Additionally, these HRS estimates were for 2012, so may not accurately represent prevalence in 2025, especially as there is evidence that prevalence rate is decreasing over time.[26] Further, because the Rush cohorts are not population-based, mortality rates and attributable risk estimates may not be representative of the general population, though we have no a priori reason to believe that the risk of death from developing Alzheimer’s would be any different in these cohorts than the general older US population. Finally, while we provide estimates adjusted for cardiovascular disease and risk factors—factors that have been proposed to account for differences between men and women in dementia risk and burden—there is still the chance for residual confounding by variables for which we did not account.

There are a number of strengths of this study, including the innovative prospective study design that bases estimates of excess deaths due to Alzheimer’s dementia and the number of deaths from Alzheimer’s in the US upon observed hazard of mortality from incident Alzheimer’s dementia, rather than what is recorded on death certificates or other methods that may underestimate the contribution of Alzheimer’s to mortality—a bias that could be particularly problematic if the bias caused by underestimation differed by sex. Our overall follow-up rate of survivors is very high, especially in ROS and MAP, the two largest studies and we have nearly complete capture of mortality leading to high internal validity. Population-based study designs typically have four or more years between assessments. Given survival times of 2-3 years after diagnosis of incident Alzheimer’s dementia in the 85+ strata, our cohorts are much more likely to capture incident disease, whereas population-based studies will likely miss many cases that exit the population before the follow-up assessment wave. The study leveraged the strength of the Rush cohorts including annual clinical evaluations and robust standardized diagnostic criteria.

This study shows that the public health burden of Alzheimer’s dementia as measured by number and proportion of deaths is much higher for both women and men than what is reported on death certificates which report a single cause of death, and that the difference for women is much more significant. The findings add to the evidence that the burden of Alzheimer’s is heightened for women by showing the number of deaths attributable to Alzheimer’s disease for women in the U.S is likely about 350,000 making it one of the leading causes of death in women, on par with heart disease and cancer. The estimated 200,000 deaths from Alzheimer’s dementia in men is also much higher than what is reported on death certificates, but less than cancer and heart disease. Overall, our findings suggest that Alzheimer’s dementia is likely an under-recognized major public health burden for women in particular.

## Data Availability

All data produced in the present study are available upon request through requests submitted online at: https://www.radc.rush.edu/home.htm

## ACKNOWLEDGMENTS

We thank all participants of the Rush Alzheimer’s Disease Center (RADC) cohorts. RADC cohort study resources can be requested at www.radc.rush.edu. The Rush Memory and Aging Project (MAP) is supported by the National Institutes of Health (NIH) (Grant R01AG017917). The Religious Orders Study (ROS), Clinical Core, and Latino Core are all sponsored by the Rush Alzheimer’s Disease Research Center’s NIH center grant (P30AG072975). The Minority Aging Research Study (MARS) is sponsored by NIH (Grant R01AG022018). The sponsor had no involvement in study design, in the collection, analysis, and interpretation of data, in the writing of the report, or in the decision to submit the article for publication.

## CONSENT STATEMENT

All RADC cohort participants provided informed consent.

